# Comparing Active Case-Finding Strategies and Passive Case-Finding for Tuberculosis: An Umbrella Review of Current Evidence

**DOI:** 10.1101/2025.06.01.25328419

**Authors:** Sonia Menon, Leon Nshimyumukiza, Pranay Sinha

## Abstract

**Background:** Tuberculosis (TB) remains a global health challenge. Recent studies have compared active case-finding (ACF) with passive case-finding (PCF) and various ACF strategies, assessing diagnostic yield, treatment outcomes, cost-effectiveness, and overall TB control impact. However, evidence remains fragmented due to methodological heterogeneity.

**Methods:** To assess how ACF compares to PCF and to different ACF strategies in improving TB detection, treatment outcomes, and overall T control, we conducted an umbrella review of systematic reviews and meta-analyses published through March 30, 2025, evaluating ACF versus PCF and different ACF strategies across diverse settings. Key outcomes included diagnostic yield, treatment outcomes, cost-effectiveness, and epidemiological impact. Methodological quality was assessed using the AMSTAR 2 tool.

**Results:** Twelve systematic reviews and meta-analyses met the inclusion criteria, with methodological quality ranging from low to moderate. ACF may be more effective than PCF in increasing TB detection rates, especially in high-risk populations such as migrants, people living with HIV, drug users, and close contacts of TB patients. Treatment outcomes among ACF-identified cases were mixed, with concerns about pre-treatment loss to follow-up. ACF interventions were cost-effective in high-prevalence settings, though their cost-effectiveness varied depending on implementation coverage, intensity, and the screening tools. However, evidence linking ACF to sustained increases in national routine TB notifications was limited.

**Conclusion:** While ACF is effective for targeted detection in high-risk groups and often cost-effective in high-burden settings, its broader epidemiological impact remains uncertain. Future TB control strategies should prioritize integrated, context-sensitive approaches that combine active and passive case detection, and future research should focus on evaluating ACF interventions based on their ability to achieve sustained reductions in TB prevalence and transmission over time.

- Take home message:
- Despite extensive research on ACF versus PCF in TB control, the fragmented and methodologically diverse evidence necessitates a comprehensive synthesis to inform global policies and practices.
- What this study adds - ACF improves TB detection in high-risk groups, but its effectiveness and cost-effectiveness are context-dependent. Cost-effective screening may enable early detection and treatment, reducing the TB burden on healthcare systems.
- ACF may be effective and cost effective in high-risk populations and at observational level but its epidemiological impact at national level is unclear
- How this study might affect research, practice or policy - This study highlights the need for targeted TB screening in high-risk groups to reduce TB incidence, support for continued investment in cost-effective strategies, and the enhancement of health outcomes through integrated approaches and supportive measures.

## Background

Tuberculosis (TB) remains a significant global health challenge, causing substantial morbidity and mortality. In 2023, 10.6 million were estimated to have TB, of whom only 7.5 million were diagnosed; approximately 1.3 million died from it, making it the second leading cause of death from a single infectious agent after coronavirus [1]. Those with socioeconomic deprivations are both at increased risk of TB disease while simultaneously having poor access to TB care Vulnerable populations include those at a disadvantaged socioeconomic position, at a higher risk of TB infection or progression to disease, and with poor access to TB care[2].

Effective treatment is crucial in managing TB and is a cornerstone of the World Health Organization’s (WHO) TB control strategy. This strategy emphasizes prompt diagnosis, effective treatment regimens, and comprehensive care to reduce transmission and prevent the emergence of drug-resistant strains. Building on the 27 % decline in TB incidence from 2000 to 2021, the WHO adopted the “END TB” strategy, which aims to reduce the number of deaths by 95 % and the incidence of TB by 90 % by 2035[3].

However, the success of treatment heavily relies on early detection of the disease, making case-finding an essential component of TB control efforts. Case-finding strategies for tuberculosis (TB) are categorized into passive case-finding (PCF) and active case-finding (ACF). PCF, the standard method in many countries, is patient-initiated and entails seeking treatment for TB symptoms[4]. This approach may miss cases, especially in high-risk groups. In contrast, ACF is a proactive, resource-intensive approach that systematically targets high-risk populations, addressing unmet needs for TB services[5]. By detecting and diagnosing TB cases early, particularly among asymptomatic individuals or those with limited access to healthcare, ACF can reduce transmission and improve treatment outcomes [6]. ACF encompasses various proactive measures beyond screening and can be tailored to identify TB in asymptomatic populations based on assessed risk [7].

Recent studies have extensively explored the relative merits of ACF versus PCF, as well as various ACF strategies, examining key outcomes such as diagnostic yield, treatment outcomes, cost-effectiveness, and overall impact on TB control. Despite the wealth of research, the evidence remains fragmented, with studies exhibiting considerable methodological heterogeneity. Understanding the strengths and limitations of each approach is crucial for informing global policies and practices in TB control efforts. Umbrella reviews, which are a synthesis of existing systematic reviews that provides top-tier evidence, can help us capture the scope of the literature [8]. Specifically, this umbrella review aims to systematically synthesize and critically appraise the existing evidence on the comparative effectiveness of ACF compared to PCF or various ACF strategies for TB detection across various settings and populations. In addition to providing an overview of the current evidence base, our review will assess the quality and strength of the evidence and identify systematic reviews with the strongest evidence.

### Methodology

This umbrella review was conducted in accordance with the Preferred Reporting Items for Systematic Reviews and Meta-Analysis (PRISMA) guidelines[9] to ensure comprehensive and transparent reporting of our review process and findings.

### Protocol Registration

The review protocol was registered in the International Prospective Register of Systematic Reviews (PROSPERO) under the identifier PROSPERO. The review protocol was registered in the International Prospective Register of Systematic Reviews (PROSPERO) under the identifier PROSPERO 2024 CRD42024514313.

Patients were not involved in the design, conduct, reporting, or dissemination plans of this umbrella review.

### Participants/population, intervention, context, and outcomes

The targeted population of our review included the general population and at-risk groups for tuberculosis. ACF was considered as the intervention and was compared to other case finding strategies, including passive case finding (PCF) as the control interventions in all contexts. All TB related clinical outcomes such as the case detection rate ratio, treatment completion rate ratio (treatment completed only), treatment success rate ratio (cure and treatment completed), and economic outcomes (e.g., incremental cost effectiveness ratios) derived from the articles rather than a predefined framework were considered.

### Search Strategy

We conducted a comprehensive literature search using four electronic databases on March 23^rd^, 2025: PubMed/MEDLINE, Embase, Scopus, and the Cochrane Library. Our search strategy combined Medical Subject Headings (MeSH) terms and relevant keywords related to tuberculosis, case-finding strategies, and specific filters for systematic reviews and meta-analyses. The detailed search strings were developed in collaboration with a research librarian to ensure sensitivity and specificity.

### Study Selection Process

Two independent reviewers (SM and LN) screened the titles and abstracts of all retrieved records to assess their relevance based on predefined inclusion and exclusion criteria. Full-text articles of potentially relevant studies were then reviewed for eligibility. Discrepancies between the reviewers were resolved through consensus or by consultation with a third reviewer (PS) to ensure objective and unbiased selection.

### Inclusion Criteria

We included studies that met the following criteria: systematic reviews with or without meta-analyses, and studies comparing ACF and PCF or different ACF strategies within diverse settings for TB detection; studies reporting on outcomes such as diagnostic yield, treatment initiation rates, treatment completion rates, cost-effectiveness, and impact on TB control measures; and studies published in peer-reviewed journals in English.

We excluded studies that did not directly compare ACF and PCF strategies or different ACF strategies for TB detection and focused exclusively on either ACF or PCF without providing comparative analysis.

### Data Extraction and Synthesis

Data extraction was independently performed by two reviewers using a standardized data extraction form. Extracted data included study characteristics (e.g., study design, population characteristics), intervention details (e.g., type of ACF/PCF strategy), and outcomes of interest (e.g., diagnostic yield, treatment initiation and completion rates, cost-effectiveness). Qualitative data from included studies were synthesized thematically to identify patterns, trends, and areas of consensus or controversy. Quantitative data from systematic reviews and meta-analyses were synthesized, and findings were presented narratively and/or descriptively, with tables and figures used to summarize key results. Key findings were stratified into specific categories, derived from the articles rather than using a predefined framework.

Given high heterogeneity in the designs, study questions, and outcomes, a descriptive presentation of relative effect estimates of meta-analyses was performed.

### Patient and Public Involvement

Patients and the public were not involved in the design, conduct, reporting, or dissemination plans of this research.

### Methodological Quality Evaluation

The methodological quality of the systematic reviews and meta-analyses included in this study was assessed using the AMSTAR 2 tool [10], which consists of 16 items designed to evaluate the methodological rigor of systematic reviews.

## Results

### Study selection

The study selection process according to PRISMA requirements [9] is summarized in Fig. 1(flow diagram). The database search retrieved a total of 217 records. After de-duplication, 112 articles were screened by title and abstract. Of these, 48 records were excluded, and 61 records were included for full text screening. Subsequently, 64 records were excluded after full text screening and 28 were assessed for eligibility. The final number of articles included was 12.

**Figure 1:**
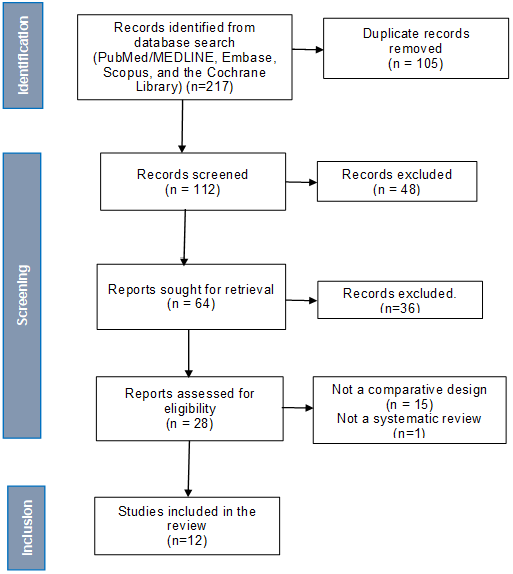
Prisma flow diagram.

### Review characteristics

Table 1 presents the characteristics of the included studies. Included reviews were meta-analyses (n=5)[11–15], systematic reviews (n=5)[16–20] and 2 systematic reviews of economic evaluations [21, 22]. All systematic reviews and meta-analyses considered Randomized controlled trials and observational studies. Various active case-finding interventions were compared to passive case detection, except in one study [13] where prevalence rates retrieved in ACF studies were compared to ones published by the WHO. All reviews considered studies without geographic restrictions and settings, except one focusing on Ethiopia [14]. Two reviews considered all TB settings (high, medium, moderate, and low)[13, 19, 20, 22], 5 reviews focused only on high TB settings [11, 12, 14, 15, 21], 2 included high and low settings [16, 17], and one considered only a low TB setting [18]. Four reviews included general and specific high-risk populations [13, 16, 17, 19], whereas other reviews considered only specific high-risk populations (people with diabetes, PLHIV, those with other respiratory diseases, cough > 2 weeks, homeless populations, and refugees)[11, 12, 14, 15, 18, 20–22]. Only 3 studies out of the 12 included reviews, 5 conducted data pooling and meta-analyses, [11–15] whereas 7 reviews provided a narrative synthesis without meta-analysis [16–19, 21].

**Table 1.**
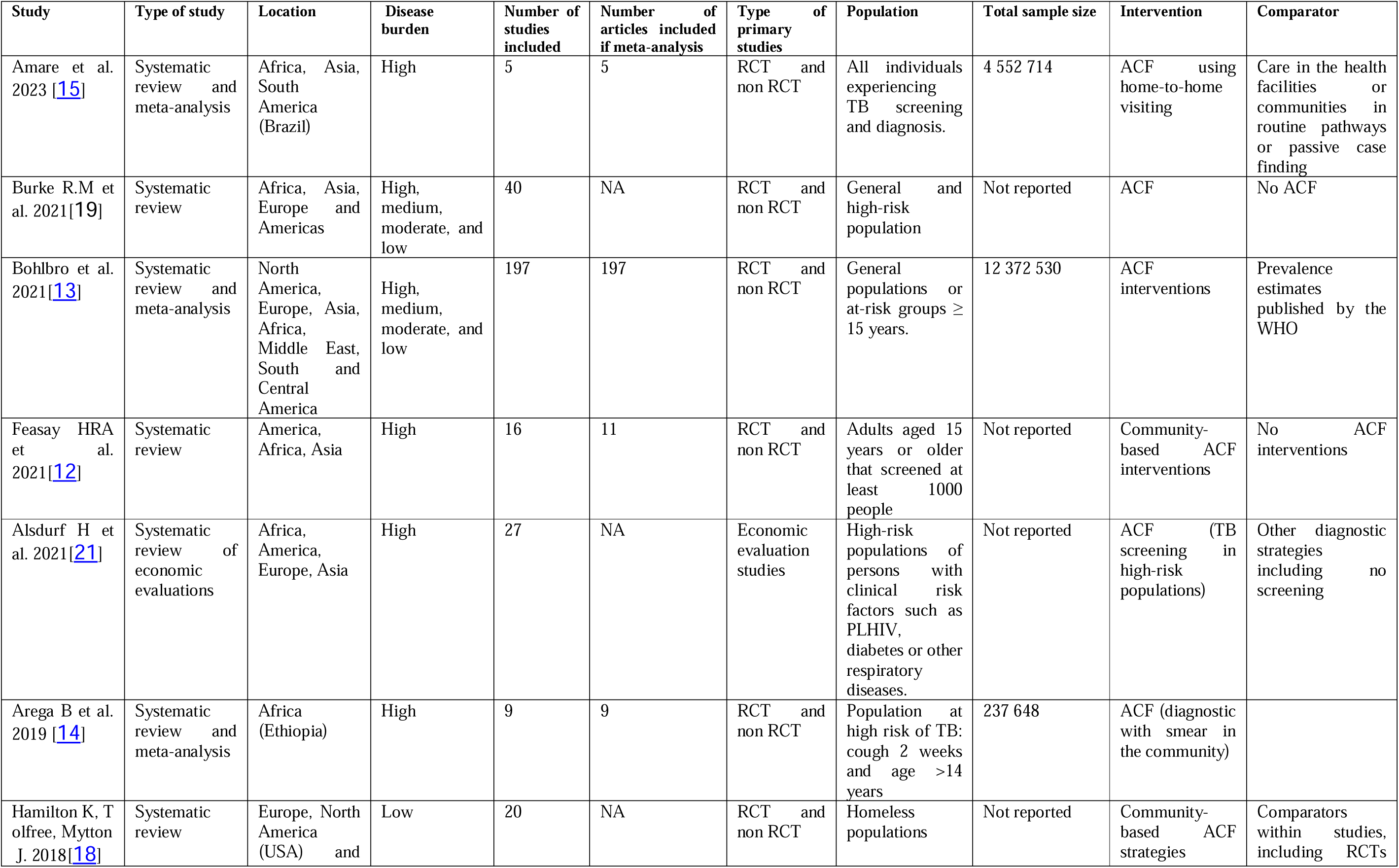
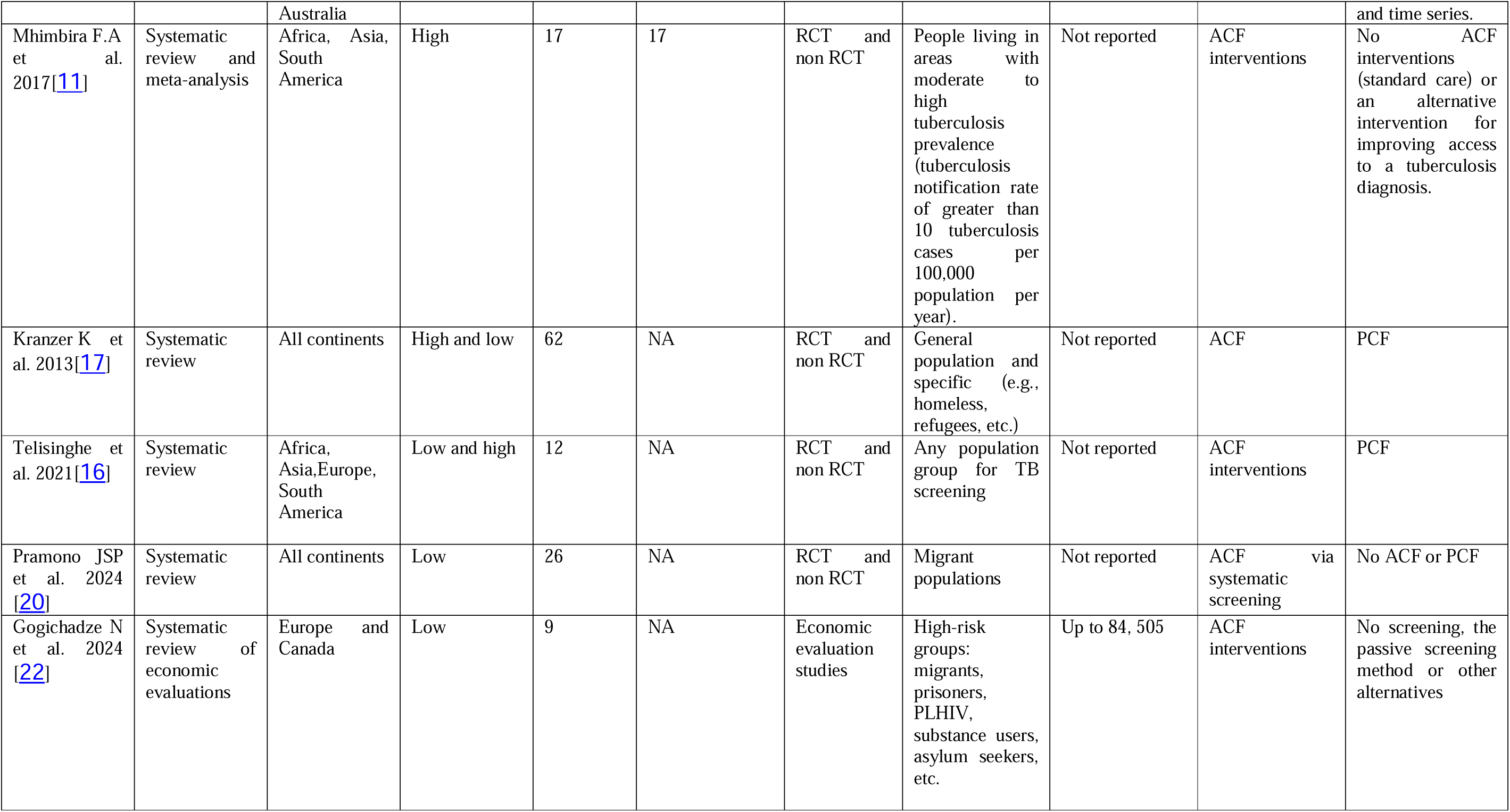
Characteristics of included studies.

### Study findings

The results summarized in Table 2 of the umbrella review on active case finding (ACF) for tuberculosis (TB) highlight several key findings across various studies regarding TB detection, case notification rates, TB treatment outcomes, and cost-effectiveness of different ACF strategies compared to passive case finding (PCF).

**Table 2.**
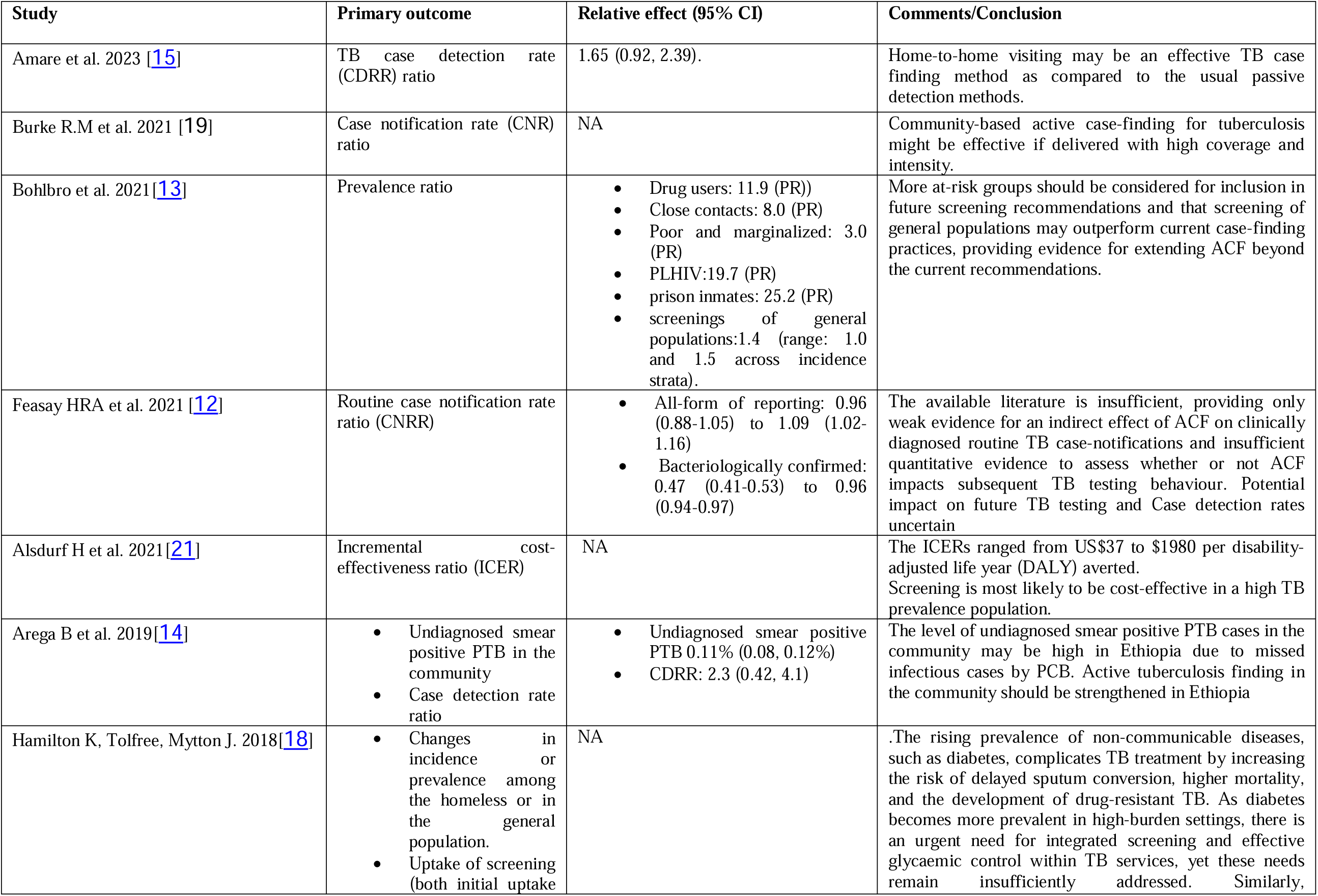
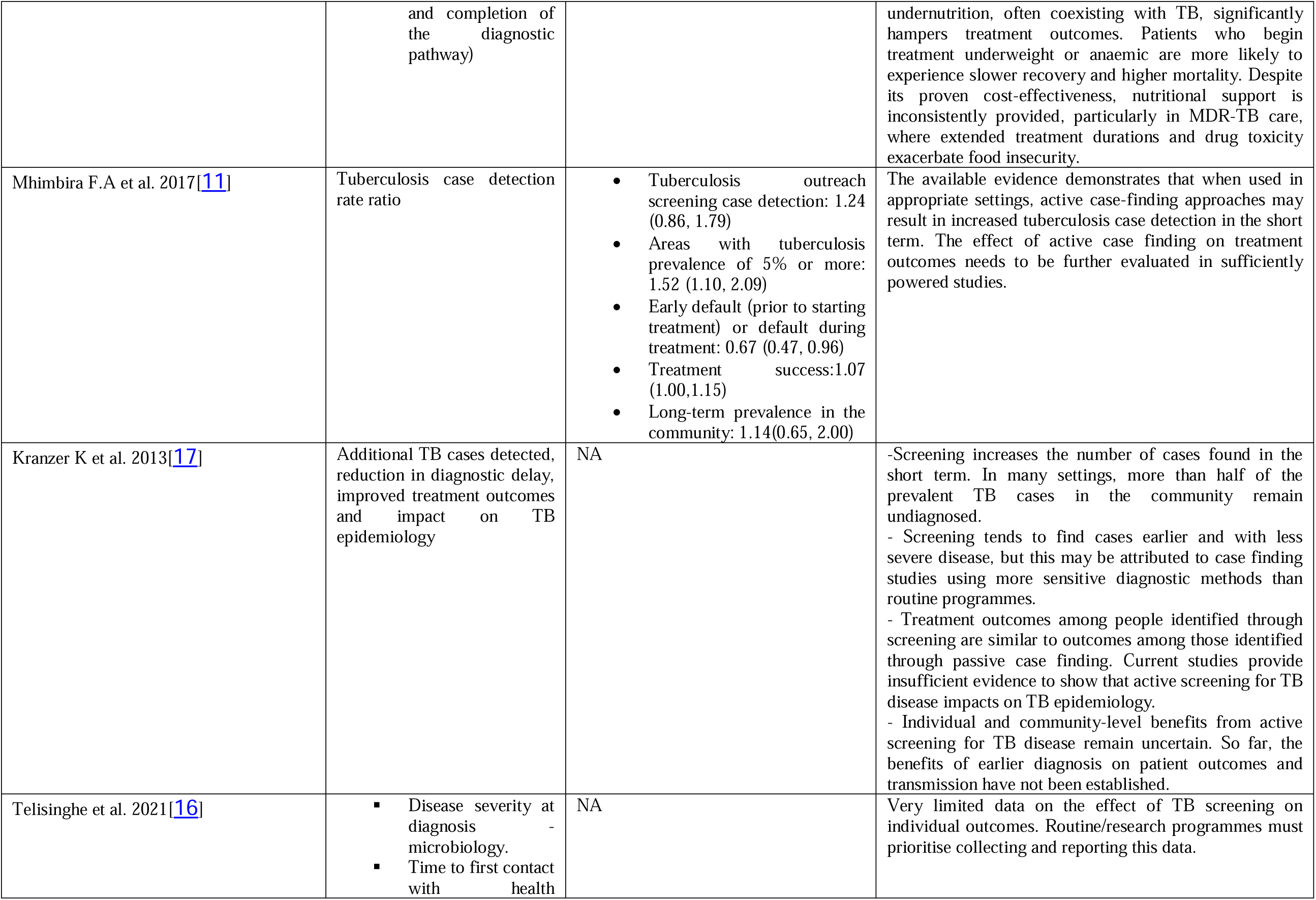
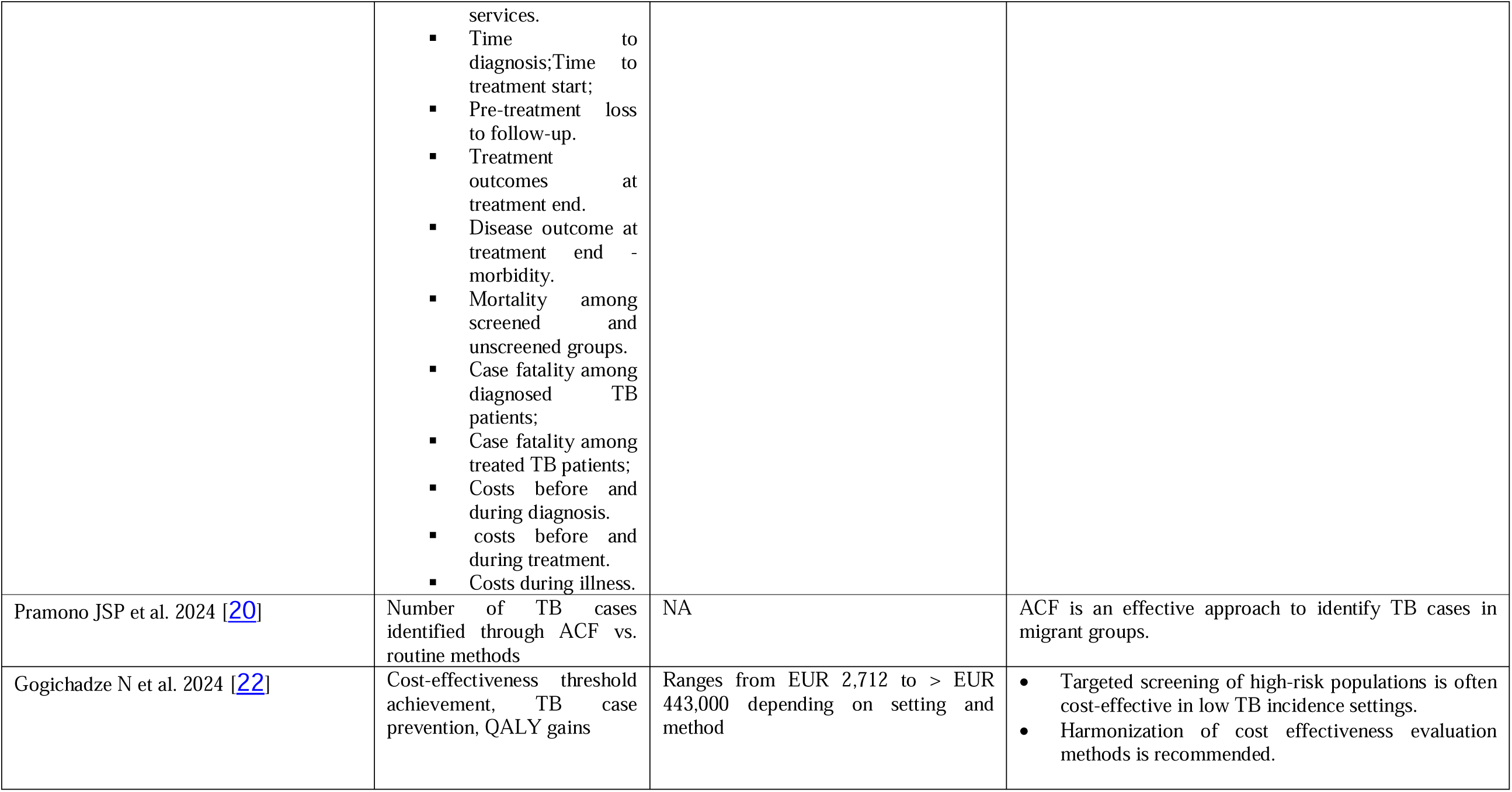
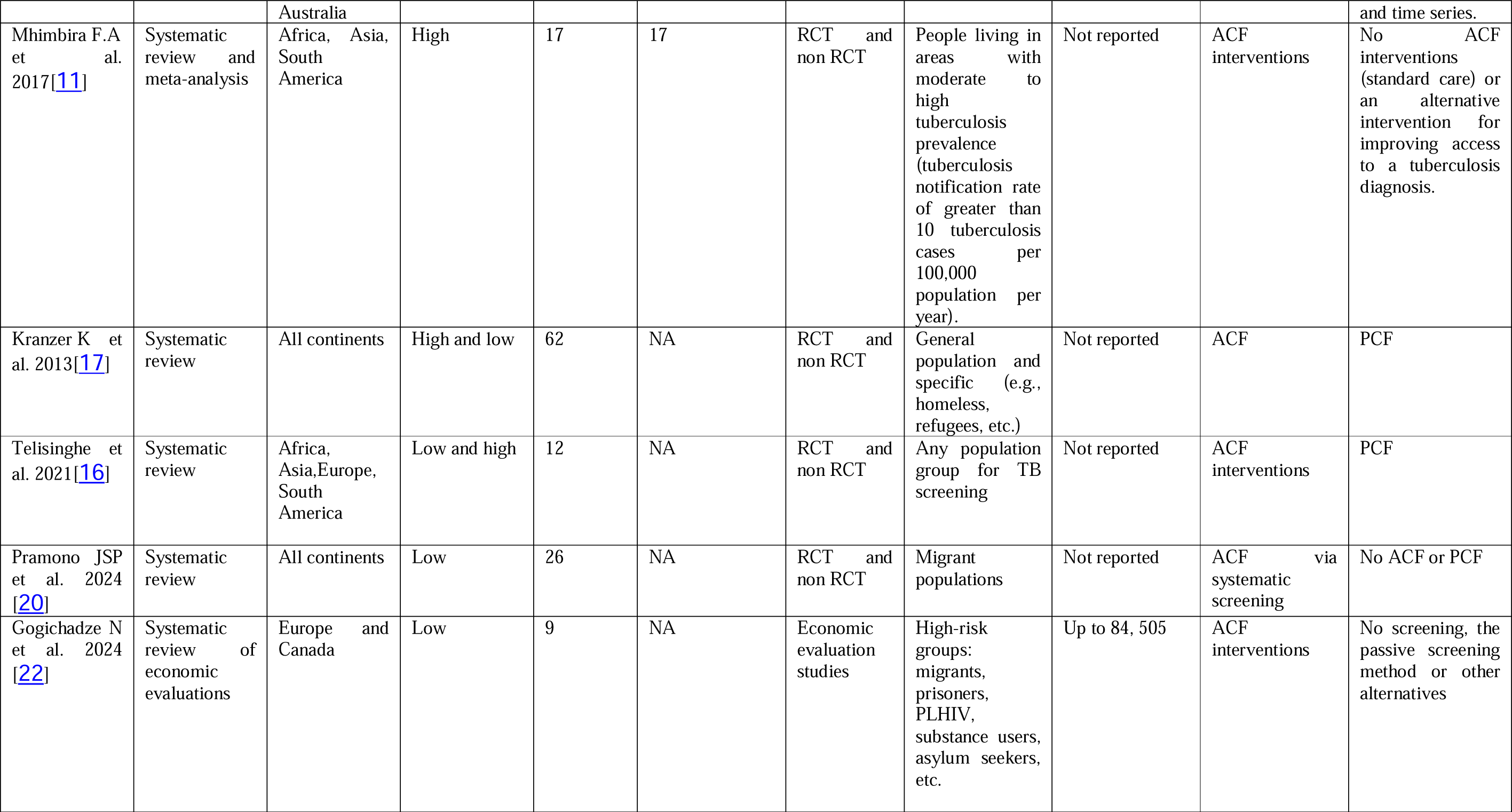
Table 2. Extraction results.

### Effectiveness of ACF versus PCF

Amare et al. (2023) [15] reported that home-to-home visits for ACF may be more effective than passive detection methods with a TB case detection rate (TCR) ratio of 1.65 (0.92 - 2.39). Burke et al. (2021)[19] found that community-based ACF was potentially effective if delivered with high coverage and intensity. Feasay et al. (2021)[12] reported weak evidence for the indirect effect of ACF on routine TB case notifications, with routine case notification rate ratios (CNRR) ranging from 0.96 (0.88-1.05) to 1.09 (1.02-1.16) for all-form reporting, and from 0.47 (0.41-0.53) to 0.96 (0.94-0.97) for bacteriologically confirmed cases. Arega et al. (2019)[14] identified a high level of undiagnosed smear-positive PTB in Ethiopia with an undiagnosed rate of 0.11% (0.08 - 0.12%) and a case detection rate ratio (CDRR) of 2.3 (0.42 - 4.1), recommending strengthened active TB finding in the community.

### ACF in TB diagnosis (both clinically and bacteriologically)

Feasay et al. (2021) reported weak evidence for the indirect effect of ACF on routine TB case notifications. Routine case notification rate ratios (CNRR) ranged from 0.96 to 1.09 for all-form reporting and from 0.47 to 0.96 for bacteriologically confirmed cases. Amare et al. (2023) found that home-to-home visits for ACF might be more effective than passive detection methods, with a TB case detection rate (TCR) ratio of 1.65 (0.92 - 2.39).

### ACF in at-risk groups

Bohlbro et al. (2021)[13] reported significant prevalence ratios (prevalence among the at-risk population compared to the general population) among various at-risk groups such as drug users (11.9), close contacts (8.0), and PLHIV (19.7), suggesting that including more at-risk groups in future screening recommendations could be beneficial. Pramono et al. (2024) [20] found ACF to be an effective approach for identifying TB cases specifically in migrant populations, an often underserved and mobile group, emphasizing the importance of tailored interventions for these high-risk settings.

### Cost-Effectiveness of ACF

Alsdurf H et al. (2021)[21] found that TB screening is likely cost-effective in high TB prevalence populations, particularly among persons living with HIV in sub-Saharan Africa among whom the incremental cost-effectiveness ratio (ICER) was as low as $37 per disability-adjusted life year (DALY) averted when the WHO four symptom screen was combined with PCR testing. However, cost-effectiveness was not clearly demonstrated among all populations with structural risk factors. For instance, a van-based screening program for persons experiencing homelessness in London had an ICER $9837 per quality-adjusted life year (QALY) gained. In Russia, case-finding among incarcerated individuals had an ICER of of $793 per TB case diagnosed. Screening in Urban slums appears to be cost-effective, with ICERs of $268 per TB case diagnosed in Cambodia. The range of published ICERs is broad because of the wide variety of testing modalities and populations studied which should caution against broad assumptions that active case-finding is universally cost-effective. Gogichadze et al. (2024) [22]further highlighted the variability in ACF cost-effectiveness, reporting thresholds ranging from EUR 2,712 to over EUR 443,000 depending on setting and method. Their study supports targeted screening as often cost-effective in low-incidence settings and calls for greater harmonization in cost-effectiveness methodologies.

### Screening uptake and effectiveness

Hamilton, Tolfree, and Mytton (2018)[18] noted that ACF appears to be effective on an observational level but emphasized the need for high-quality evidence to optimize programs. They suggested that incentives and professional support could improve screening uptake and completion. Mhimbira et al. (2017)[11] reported that ACF increases TB case detection in the short term, especially with sensitive diagnostic methods. However, they found insufficient evidence of the long-term benefits and impact on TB epidemiology. Telisinghe et al. (2021)[16] examined various measures of TB screening impact, including disease severity, time to diagnosis, and costs, emphasizing the need for more data on individual outcomes, but found limited evidence to show milder disease and better treatment outcomes in ACF as compared to PCF. They recommended prioritizing data collection in routine and research programs to better understand the effects of active TB screening.

### ACF and treatment outcomes

Mhimbira et al. (2017) [11] found that ACF may improve treatment success rates but also noted an increased early default rate prior to or during treatment. According to Kranzer et al. (2013) [17], ACF can identify more TB cases in the short term, but the treatment outcomes among those identified through ACF may not be superior to those identified through PCF. The benefits of earlier diagnosis on patient outcomes and transmission remain uncertain, making the overall individual and community-level benefits of ACF unclear. Telisinghe et al. (2021) [16] examined various metrics such as disease severity at diagnosis, time to first contact with health services, time to diagnosis and treatment start, pre-treatment loss to follow-up, and treatment outcomes at the end of treatment. They found that while ACF can improve detection rates, its broader impact on treatment outcomes and TB control remains uncertain.

### Methodological quality

Figure 2 Risk of bias summarizes the frequency of each AMSTAR2 rating for each domain across reviews. Overall, the includedreviews were generally of low to moderate methodological quality. This rating was primarily the lack of a list of excluded studies (item 7), and insufficient information regarding the source of funding in the included studies (item 10). The absence of a comprehensive list of excluded studies in many reviews (9 out of 12) limited the transparency and replicability of these reviews. Poor reporting of funding sources (7 out of 12) in the primary included studies raised concerns about potential conflicts of interest and the influence of funding on study outcomes.

**Figure 2:**
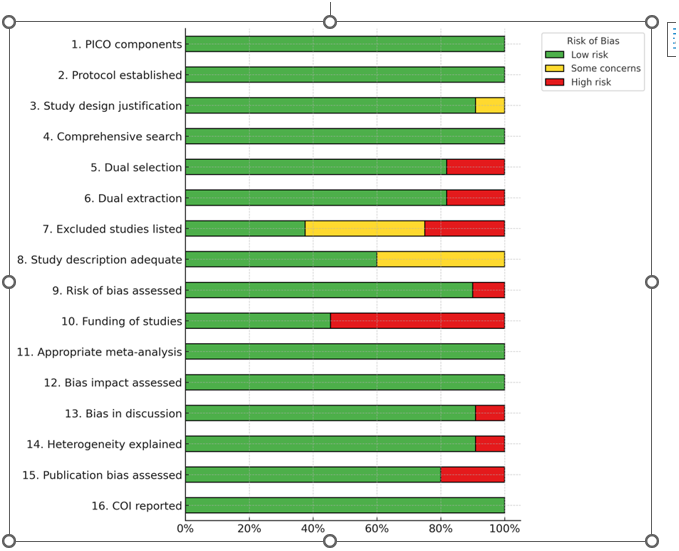
Risk of bias.

## Discussion

The current umbrella review summarized and systematically appraised the evidence about the effectiveness and cost-effectiveness of ACF compared to other strategies including PCF for TB detection. The summary message provided by the 12 systematic reviews and meta-analyses is that the evidence about the effectiveness and cost-effectiveness of TB ACF remains limited, context-dependent and related to various factors such as coverage, intensity, and screening tools used Regarding the effectiveness, this umbrella review shows that ACF may be effective in high-risk groups, such as people living with HIV, close contacts of TB patients, and individuals in high-prevalence areas such as urban slums. Indeed, targeted ACF interventions in these populations may lead to increased TB case detection, particularly for bacteriologically confirmed cases. However, the broader impact of ACF on national or population-level TB incidence remains unclear.

While ACF is supposed to boost case detection in targeted groups, these interventions do not appear to significantly alter the overall epidemiology of TB and there is limited evidence indicating a direct and positive effect of ACF on bacteriologically diagnosed routine TB case notifications. This suggests that while ACF is valuable for identifying and treating TB cases that might otherwise remain undiagnosed, it should be viewed as a complementary strategy rather than a standalone solution for TB control.

This is aligned with the findings of Koura et al. (2017) [23] who found that while ACF interventions led to substantial increases in TB case notifications within intervention areas, these gains rarely translated into significant changes at the national level. Only one country (Benin) demonstrated a statistically significant increase in national case notifications during the intervention period that could plausibly be attributed to ACF, largely because the intervention covered the entire country. The study underscores that while ACF can be impactful locally, its broader epidemiological effects may be limited without national scale-up—due in part to high costs, limited health system capacity, and challenges in sustaining intensive interventions over time.

This umbrella review highlights a persistent gap in the evidence: the link between TB detection through active case finding (ACF) and downstream treatment outcomes remains underexplored. While ACF can identify cases earlier sometimes among asymptomatic individuals the implications for treatment initiation, adherence, and completion are unclear. Research is especially needed to understand how acceptable treatment is for asymptomatic individuals, how often they are lost to follow-up before starting therapy, and what strategies improve retention under routine programmatic conditions. Ultimately, evaluating ACF’s true effectiveness requires tracking individuals from diagnosis through treatment completion.

The effectiveness of ACF is strongly influenced by implementation intensity and coverage. For example, Marks et al. (2019) conducted a cluster-randomized trial in Vietnam where the entire adult population of Ca Mau Province was screened annually using sputum Xpert testing, regardless of symptoms. [24]. TB prevalence declined by 44%, but this required screening 1,002 people per year for three years to prevent one prevalent case in the fourth year. While the study demonstrates that intensive ACF can reduce transmission, it also illustrates the labor and resource demands of high-coverage strategies, raising questions about scalability in other settings.

The choice of screening tools and delivery methods also impacts ACF effectiveness. Uncertainty remains over whether house-to-house visits outperform geographically closer mobile or outreach clinics in detecting microbiologically confirmed TB. Each approach has trade-offs in reach, logistics, and cost. Socioeconomic factors such as poverty, healthcare access, and infrastructure must inform which strategies are adopted. Comparative evaluations are needed to determine the most effective and sustainable screening modalities in different settings.

Despite limited data, this review supports the cost-effectiveness of ACF in high-burden and high-risk populations. Interventions using symptom screening combined with chest X-ray or molecular testing (e.g., Xpert) can achieve high diagnostic yield at reasonable cost, particularly when targeted to vulnerable groups. Gomes et al. (2022) assessed ACF across 20 countries in TB REACH Wave 5 and emphasized the importance of tailoring strategies to local epidemiology and resources. [25] Selecting cost-effective tools is key to enhancing the coverage and intensity of ACF without escalating costs. If ACF intervention may drive down transmission, the prevented secondary infections are likely to enhance the cost-effectiveness of these interventions.

This review supports a targeted, context-sensitive approach to ACF. Screening high-risk groups with tailored interventions can reduce TB incidence within these populations and contribute to overall TB control. Integrated strategies that combine active and passive case detection can broaden reach, particularly in high-prevalence settings. Supportive measures, including patient incentives and professional assistance may improve screening uptake and reduce pre-treatment loss to follow-up. Continued investment in cost-effective screening is justified by the potential for earlier detection and reduced TB burden. Comprehensive data collection, particularly on the long-term outcomes of TB screening in diverse contexts is essential to inform future interventions and policy decisions. Robust research can provide evidence-based insights that guide the development and implementation of effective TB elimination strategies.

Our umbrella review possesses significant strengths. It includes studies regardless of the language of publication, which reduces potential language bias. Also, we also covered several databases thus minimizing publication bias.

However, our umbrella review has several limitations that should be acknowledged. These limitations are multifaceted and impact the overall interpretation and applicability of the findings. Firstly, ten systematic reviews were included in the final analysis and the methodological quality of the included studies was generally moderate. Significant issues included the absence of protocol registration, the lack of comprehensive lists of excluded studies, and insufficient information regarding the sources of funding for the included studies. These factors introduce potential biases and reduce the overall reliability of the evidence.

Another notable limitation is the methodological heterogeneity across the included systematic reviews and meta-analyses. The diversity in study designs, populations, and outcome measures complicates direct comparisons and synthesis of the findings. This heterogeneity hampers the ability to draw definitive conclusions about the relative effectiveness and cost-effectiveness of ACF compared to PCF and various ACF strategies.

But perhaps the most significant limitation of this umbrella review reflects a broader limitation in the field itself: the included systematic reviews and the primary studies they synthesize have largely focused on intermediate outcomes such as case detection and diagnostic yield, rather than on more consequential endpoints like reductions in TB prevalence or transmission within the screened populations. These limitations highlight the need for more robust, context-specific research to fill the existing evidence gaps and provide clearer insights into the effectiveness and cost-effectiveness of ACF in different settings.

## Conclusion

This review supports a targeted, context-sensitive approach to ACF. Screening high-risk groups with tailored interventions can reduce TB incidence within these populations and contribute to overall TB control. Integrated strategies that combine active and passive case detection can broaden reach, particularly in high-prevalence settings. Supportive measures, including patient incentives and professional assistance may improve screening uptake and reduce pre-treatment loss to follow-up. Continued investment in cost-effective screening is justified by the potential for earlier detection and reduced TB burden. Comprehensive data collection, particularly on the long-term outcomes of TB screening in diverse contexts is essential to inform future interventions and policy decisions. Robust research can provide evidence-based insights that guide the development and implementation of effective TB elimination strategies.

## Supporting information

Search strategy

Search strategy

## Data Availability

All data produced in the present work are contained in the manuscript

## Declarations

### Ethics approval and consent to participate

Not applicable. This study is an umbrella review of previously published systematic reviews and does not involve human participants or collection of primary data.

### Consent for publication

Not applicable

### Availability of data and material

Yes, in the submission

### Authors’ contributions

SM and PS conceived the study; Search strategy: LN Abstract and full text screening; LN and SM ; Extraction; LN and SM; Drafting and interpretation; SM, LN, PS Validation, SM, LN,PS All authors have read and agreed to the published version of the manuscript

## Acknowledgments

We would like to acknowledge Prof. Anthony D. Harries for his efforts in reading and editing the prefinal document.

---

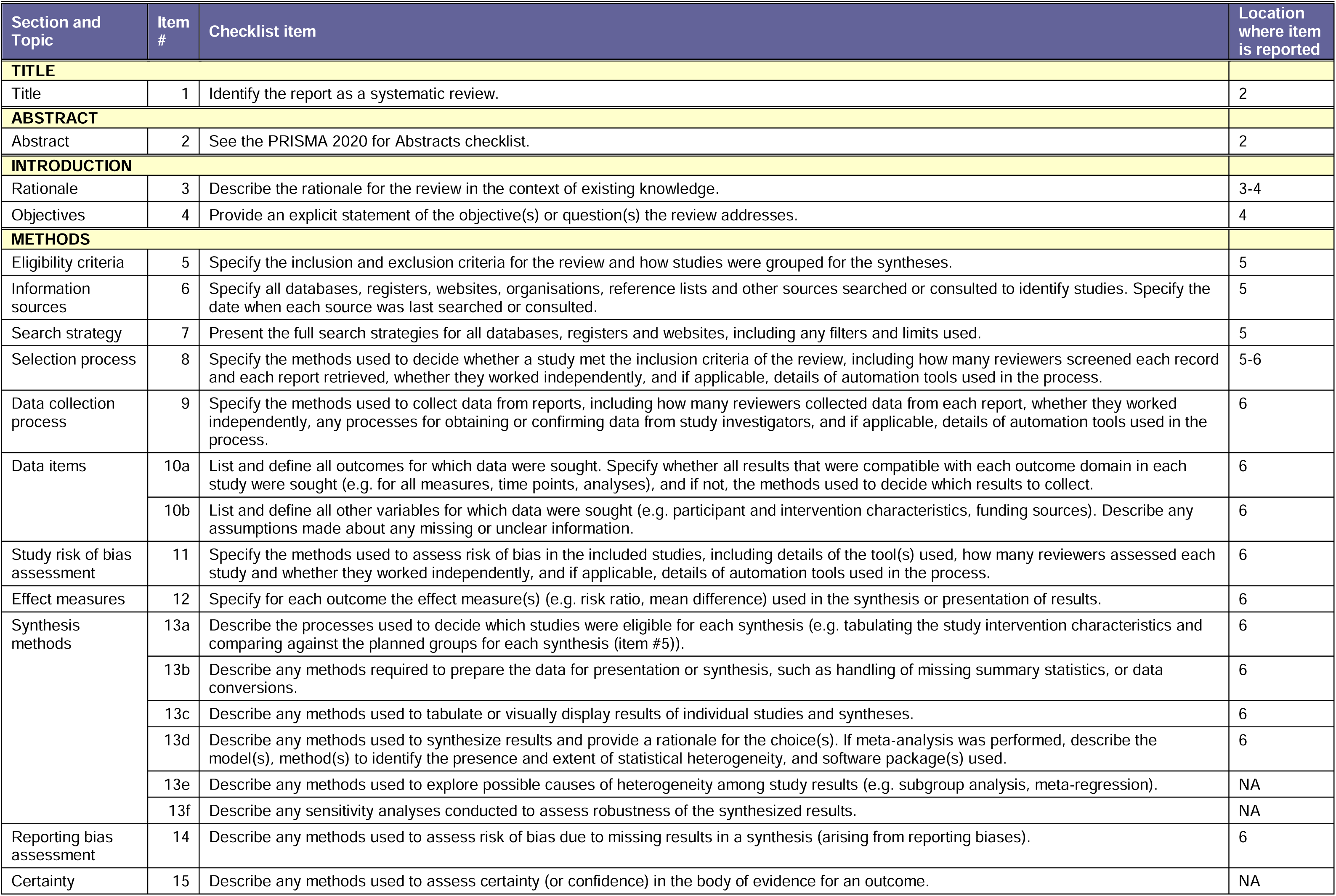
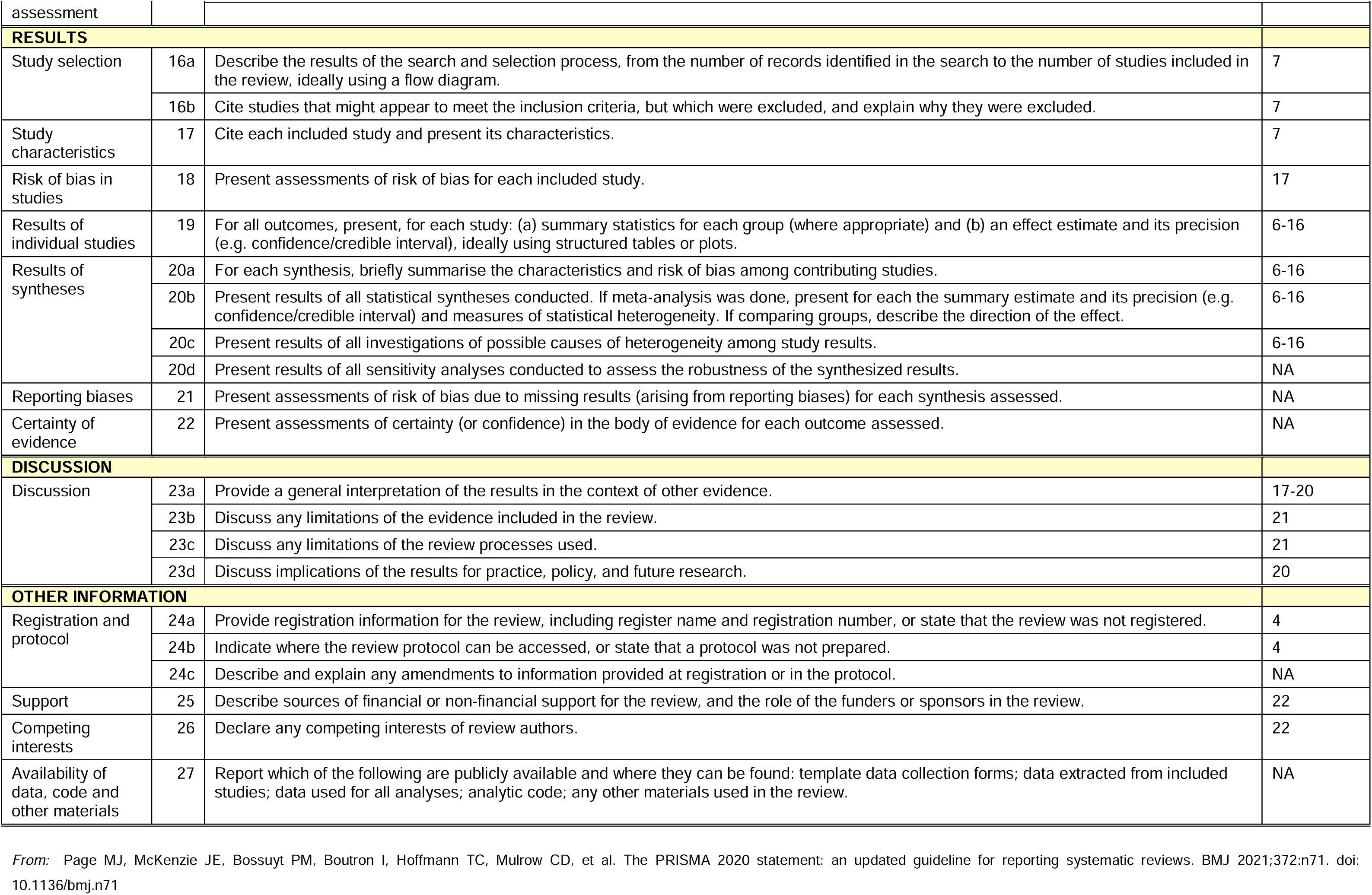

PRISMA 2020 Checklist
| Section and Topic | Item # | Checklist item | Location where item is reported |
| --- | --- | --- | --- |
| **TITLE** |  |  |  |
| Title | 1 | Identify the report as a systematic review. | 2 |
| **ABSTRACT** |  |  |  |
| Abstract | 2 | See the PRISMA 2020 for Abstracts checklist. | 2 |
| **INTRODUCTION** |  |  |  |
| Rationale | 3 | Describe the rationale for the review in the context of existing knowledge. | 3-4 |
| Objectives | 4 | Provide an explicit statement of the objective(s) or question(s) the review addresses. | 4 |
| **METHODS** |  |  |  |
| Eligibility criteria | 5 | Specify the inclusion and exclusion criteria for the review and how studies were grouped for the syntheses. | 5 |
| Information sources | 6 | Specify all databases, registers, websites, organisations, reference lists and other sources searched or consulted to identify studies. Specify the date when each source was last searched or consulted. | 5 |
| Search strategy | 7 | Present the full search strategies for all databases, registers and websites, including any filters and limits used. | 5 |
| Selection process | 8 | Specify the methods used to decide whether a study met the inclusion criteria of the review, including how many reviewers screened each record and each report retrieved, whether they worked independently, and if applicable, details of automation tools used in the process. | 5-6 |
| Data collection process | 9 | Specify the methods used to collect data from reports, including how many reviewers collected data from each report, whether they worked independently, any processes for obtaining or confirming data from study investigators, and if applicable, details of automation tools used in the process. | 6 |
| Data items | 10a | List and define all outcomes for which data were sought. Specify whether all results that were compatible with each outcome domain in each study were sought (e.g. for all measures, time points, analyses), and if not, the methods used to decide which results to collect. | 6 |
|  | 10b | List and define all other variables for which data were sought (e.g. participant and intervention characteristics, funding sources). Describe any assumptions made about any missing or unclear information. | 6 |
| Study risk of bias assessment | 11 | Specify the methods used to assess risk of bias in the included studies, including details of the tool(s) used, how many reviewers assessed each study and whether they worked independently, and if applicable, details of automation tools used in the process. | 6 |
| Effect measures | 12 | Specify for each outcome the effect measure(s) (e.g. risk ratio, mean difference) used in the synthesis or presentation of results. | 6 |
| Synthesis methods | 13a | Describe the processes used to decide which studies were eligible for each synthesis (e.g. tabulating the study intervention characteristics and comparing against the planned groups for each synthesis (item #5)). | 6 |
|  | 13b | Describe any methods required to prepare the data for presentation or synthesis, such as handling of missing summary statistics, or data conversions. | 6 |
|  | 13c | Describe any methods used to tabulate or visually display results of individual studies and syntheses. | 6 |
|  | 13d | Describe any methods used to synthesize results and provide a rationale for the choice(s). If meta-analysis was performed, describe the model(s), method(s) to identify the presence and extent of statistical heterogeneity, and software package(s) used. | 6 |
|  | 13e | Describe any methods used to explore possible causes of heterogeneity among study results (e.g. subgroup analysis, meta-regression). | NA |
|  | 13f | Describe any sensitivity analyses conducted to assess robustness of the synthesized results. | NA |
| Reporting bias assessment | 14 | Describe any methods used to assess risk of bias due to missing results in a synthesis (arising from reporting biases). | 6 |
| Certainty | 15 | Describe any methods used to assess certainty (or confidence) in the body of evidence for an outcome. | NA |

PRISMA 2020 Checklist
| Section and Topic | Item # | Checklist item | Location where item is reported |
| --- | --- | --- | --- |
| assessment |  |  |  |
| **RESULTS** |  |  |  |
| Study selection | 16a | Describe the results of the search and selection process, from the number of records identified in the search to the number of studies included in the review, ideally using a flow diagram. | 7 |
|  | 16b | Cite studies that might appear to meet the inclusion criteria, but which were excluded, and explain why they were excluded. | 7 |
| Study characteristics | 17 | Cite each included study and present its characteristics. | 7 |
| Risk of bias in studies | 18 | Present assessments of risk of bias for each included study. | 17 |
| Results of individual studies | 19 | For all outcomes, present, for each study: (a) summary statistics for each group (where appropriate) and (b) an effect estimate and its precision (e.g. confidence/credible interval), ideally using structured tables or plots. | 6-16 |
| Results of syntheses | 20a | For each synthesis, briefly summarise the characteristics and risk of bias among contributing studies. | 6-16 |
|  | 20b | Present results of all statistical syntheses conducted. If meta-analysis was done, present for each the summary estimate and its precision (e.g. confidence/credible interval) and measures of statistical heterogeneity. If comparing groups, describe the direction of the effect. | 6-16 |
|  | 20c | Present results of all investigations of possible causes of heterogeneity among study results. | 6-16 |
|  | 20d | Present results of all sensitivity analyses conducted to assess the robustness of the synthesized results. | NA |
| Reporting biases | 21 | Present assessments of risk of bias due to missing results (arising from reporting biases) for each synthesis assessed. | NA |
| Certainty of evidence | 22 | Present assessments of certainty (or confidence) in the body of evidence for each outcome assessed. | NA |
| **DISCUSSION** |  |  |  |
| Discussion | 23a | Provide a general interpretation of the results in the context of other evidence. | 17-20 |
|  | 23b | Discuss any limitations of the evidence included in the review. | 21 |
|  | 23c | Discuss any limitations of the review processes used. | 21 |
|  | 23d | Discuss implications of the results for practice, policy, and future research. | 20 |
| **OTHER INFORMATION** |  |  |  |
| Registration and protocol | 24a | Provide registration information for the review, including register name and registration number, or state that the review was not registered. | 4 |
|  | 24b | Indicate where the review protocol can be accessed, or state that a protocol was not prepared. | 4 |
|  | 24c | Describe and explain any amendments to information provided at registration or in the protocol. | NA |
| Support | 25 | Describe sources of financial or non-financial support for the review, and the role of the funders or sponsors in the review. | 22 |
| Competing interests | 26 | Declare any competing interests of review authors. | 22 |
| Availability of data, code and other materials | 27 | Report which of the following are publicly available and where they can be found: template data collection forms; data extracted from included studies; data used for all analyses; analytic code; any other materials used in the review. | NA |

