## Supplementary material for "Comparing Active Case-Finding Strategies and Passive Case-Finding for Tuberculosis: An Umbrella Review of Current Evidence": Search strategy

**TB umbrella review- Search strategy**

### 1. PubMed

("tuberculosis"[MeSH Terms] OR tuberculosis[tiab] OR TB[tiab])
AND
(
 "case finding"[tiab] OR "case detection"[tiab] OR "screening"[tiab] OR
 "active case finding"[tiab] OR ACF[tiab] OR "active TB screening"[tiab] OR "systematic screening"[tiab] OR
 "passive case finding"[tiab] OR PCF[tiab] OR "routine care"[tiab] OR "no screening"[tiab] OR "standard care"[tiab] OR "usual care"[tiab]
)
AND
(
 "effectiveness"[tiab] OR "impact"[tiab] OR "benefit"[tiab] OR "value"[tiab] OR
 "yield"[tiab] OR "diagnostic yield"[tiab] OR "screening yield"[tiab] OR "TB yield"[tiab] OR
 "case yield"[tiab] OR "detection rate"[tiab] OR "case detection rate"[tiab] OR
 "diagnosed cases"[tiab] OR "cases detected"[tiab] OR "missed cases"[tiab] OR
 "true positives"[tiab] OR "prevalence detected"[tiab] OR "test positivity"[tiab] OR
 "number needed to screen"[tiab] OR
 "cost effectiveness"[tiab] OR "cost-effectiveness"[tiab] OR
 "cost efficiency"[tiab] OR "cost-efficiency"[tiab] OR
 "cost benefit"[tiab] OR "cost-benefit"[tiab] OR
 "cost utility"[tiab] OR "cost-utility"[tiab] OR
 "economic evaluation"[tiab] OR "economic analysis"[tiab] OR
 "health economics"[tiab] OR "cost analysis"[tiab] OR
 "resource use"[tiab] OR "cost outcome"[tiab] OR "value for money"[tiab] OR "budget impact"[tiab]
)
AND
(
 "systematic review"[Publication Type] OR "meta-analysis"[Publication Type] OR
 "systematic review"[tiab] OR "meta-analysis"[tiab] OR "review"[tiab]
)
NOT
(
 "letter"[Publication Type] OR "comment"[Publication Type] OR "editorial"[Publication Type]
)

### 2. Embase (via Ovid ou Embase.com)

('tuberculosis'/exp OR tuberculosis:ti,ab OR tb:ti,ab)
AND
(
 'case finding':ti,ab OR 'case detection':ti,ab OR screening:ti,ab OR
 'active case finding':ti,ab OR acf:ti,ab OR 'active tb screening':ti,ab OR 'systematic screening':ti,ab OR
 'passive case finding':ti,ab OR pcf:ti,ab OR 'routine care':ti,ab OR 'no screening':ti,ab OR 'standard care':ti,ab OR 'usual care':ti,ab
)
AND
(
 effectiveness:ti,ab OR impact:ti,ab OR benefit:ti,ab OR value:ti,ab OR
 yield:ti,ab OR 'diagnostic yield':ti,ab OR 'screening yield':ti,ab OR 'tb yield':ti,ab OR
 'case yield':ti,ab OR 'detection rate':ti,ab OR 'case detection rate':ti,ab OR
 'diagnosed cases':ti,ab OR 'cases detected':ti,ab OR 'missed cases':ti,ab OR
 'true positives':ti,ab OR 'prevalence detected':ti,ab OR 'test positivity':ti,ab OR
 'number needed to screen':ti,ab OR
 'cost effectiveness':ti,ab OR 'cost-effectiveness':ti,ab OR
 'cost efficiency':ti,ab OR 'cost-efficiency':ti,ab OR
 'cost benefit':ti,ab OR 'cost-benefit':ti,ab OR
 'cost utility':ti,ab OR 'cost-utility':ti,ab OR
 'economic evaluation':ti,ab OR 'economic analysis':ti,ab OR
 'health economics':ti,ab OR 'cost analysis':ti,ab OR
 'resource use':ti,ab OR 'cost outcome':ti,ab OR 'value for money':ti,ab OR 'budget impact':ti,ab
)
AND
(
 'systematic review'/exp OR 'meta analysis'/exp OR
 'systematic review':ti,ab OR 'meta-analysis':ti,ab OR review:ti,ab
)
NOT
(
 'conference abstract'/it OR 'editorial'/it OR 'letter'/it
)

### 3. Cochrane Library

(tuberculosis OR TB)
AND
(
 "case finding" OR "case detection" OR screening OR
 "active case finding" OR ACF OR "active TB screening" OR "systematic screening" OR
 "passive case finding" OR PCF OR "routine care" OR "no screening" OR "standard care" OR "usual care"
)
AND
(
 effectiveness OR impact OR benefit OR value OR
 yield OR "diagnostic yield" OR "screening yield" OR "TB yield" OR
 "case yield" OR "detection rate" OR "case detection rate" OR
 "diagnosed cases" OR "cases detected" OR "missed cases" OR
 "true positives" OR "prevalence detected" OR "test positivity" OR
 "number needed to screen" OR
 "cost effectiveness" OR "cost-effectiveness" OR
 "cost efficiency" OR "cost-efficiency" OR
 "cost benefit" OR "cost-benefit" OR
 "cost utility" OR "cost-utility" OR
 "economic evaluation" OR "economic analysis" OR
 "health economics" OR "cost analysis" OR
 "resource use" OR "cost outcome" OR "value for money" OR "budget impact"
)
AND
(
 "systematic review" OR "meta-analysis" OR review
)
