## Supplementary material for "Comparing Active Case-Finding Strategies and Passive Case-Finding for Tuberculosis: An Umbrella Review of Current Evidence": Search strategy

### Umbrella review TB: search strategy (1946 to March 23^rd^, 2025)

| **N°** | **Search** |
| --- | --- |
| 1 | tuberculosis/ OR lung tuberculosis/ |
| 2 | ‘Tuberculosis OR TB OR Mycobacterium |
| 3 | 1 OR 2 |
| 4 | ‘Active case finding’ or ‘ACF’ [Title/Abstract] |
| 5 | 3 AND 4 |
| 6 | ("meta analysis as topic"[MeSH Terms]) OR ("systematic reviews as topic"[MeSH Terms]) OR ("meta analysis"[Title/Abstract]) OR ("systematic review"[Title/Abstract]) |
| 7 | 5 AND 6 |
| 8 | Case Reports/ OR Case Reports as Topic/ OR Comment/ OR Editorial/ OR Letter/ OR Overall/ OR Single-Case Studies as Topic/ |
| 9 | ((case ADJ (report OR series OR study)) OR conference OR congress OR editor* OR errata OR erratum OR letter* OR meeting abstract OR reply OR replies OR "structured abstract").ti. |
| 10 | 8 OR 9 |
| 11 | 7 NOT 10 |
| 12 | exp Budgets/ OR exp "Costs AND Cost Analysis"/ OR exp Decision Theory/ OR Economics/ OR Economics, Dental/ OR exp Economics, Hospital/ OR Economics, Medical/ OR Economics, Nursing/ OR Economics, Pharmaceutical/ OR exp "Fees AND Charges"/ OR Markov Chains/ OR exp Models, Economic/ OR Monte Carlo Method/ |
| 13 | (cost OR costing OR costly OR costs OR economic* OR expenditure OR expenditures OR expense OR expenses OR finance OR financed OR finances OR financial OR pharmacoeconomic* OR pharmaco-economic* OR price OR prices OR pricing).ti,bt,kf. |
| 14 | (cost OR costing OR costly OR costs OR economic* OR expenditure OR expenditures OR expense OR expenses OR finance OR financed OR finances OR financial OR pharmacoeconomic* OR pharmaco-economic* OR price OR prices OR pricing).ab. /freq=2 |
| 15 | (cost* ADJ2 (analy* OR benefit* OR effective* OR efficacy OR efficien* OR minimi* OR outcome OR outcomes OR utilit*)).ab,kf. |
| 16 | (value ADJ2 (monetary OR money)).ti,bt,ab,kf. |
| 17 | economic model*.ab,kf. |
| 18 | ((decision* ADJ2 (analy* OR model* OR tree*)) OR markov OR monte carlo).ti,bt,ab,kf. |
| 19 | OR/16-23 |
| 20 | 19 AND 5 |
| 21 | 11 OR 20 |
